# Assessment of Quadriceps Muscle Weakness in Association with Symptomatic and Radiological Osteoarthritis of the Knee

**DOI:** 10.1101/2021.07.30.21261384

**Authors:** Siddhi Hegde, N Ranganath

## Abstract

**Background:** Primary knee osteoarthritis is a significant cause of impairment of the lower limb’s mobility, making effective rehabilitation particularly important. This study aimed to identify the association of quadriceps strength with pain and disability in comparison to its association with increasing severity of radiological grades of knee osteoarthritis.

**Methods:** 50 subjects above the age of 40 years with different grades of knee osteoarthritis were evaluated for their radiological grade of osteoarthritis using Kellgren-Lawrence grading criteria for the knee radiographs, isometric quadriceps muscle strength using a modified hand-held dynamometer, pain, and disability using WOMAC questionnaire, all on the same day in this cross-sectional study.

**Results:** There is a significant negative correlation between K/L grade and maximum strength of the quadriceps muscle, r **(50) = -0**.**28** (p<0.05). We found a highly significant negative correlation between WOMAC score and strength of the quadriceps muscle, **r (50) = -0**.**41** (p<0.05).

**Discussion:** Quadriceps strength reduced progressively in subjects with more significant radiographic changes, questioning whether quadriceps muscle weakness precedes the onset of osteoarthritis and progressively results in further damage or osteoarthritis itself reduces the quadriceps strength. Though patients may have greater joint destruction on knee radiographs, increasing the strength of quadriceps might reduce the pain and functional disability.

## INTRODUCTION

The knee has enjoyed much scientific attention due to its essential role in transmission, absorption, and redistribution of forces during daily activities. The healthy knee allows for joint stability in ambulation across different terrains. Osteoarthritis is the most common chronic joint disease, which involves the deterioration of joint cartilage and subjacent bone ^[1]^. It is marked by cell stress and extracellular matrix deterioration initiated by micro- and macro-trauma that establish dysfunctional repair pathways, including pro-inflammatory pathways of innate immunity ^[2]^.

Primary knee osteoarthritis significantly impedes the mobility and stability of the lower limb. There is no known cure for this evolving degenerative disease, making effective rehabilitation specifically necessary ^[3]^.

In persons at risk, a change in modifiable risk factors such as repetitive movements of joints, obesity, infection, and injuries, amongst many others, may reduce their susceptibility to the disease ^[3]^. Quadriceps muscle strength has been shown to influence the lower limbs’ pain and disability, apart from other pre-established factors influencing knee osteoarthritis. A vital clinical indicator of knee osteoarthritis is knee pain ^[4]^. Symptomatic knee osteoarthritis is defined as a Kellgren-Lawrence grade of at least mild radiographic osteoarthritis and symptoms in the same knee. It is important in clinical diagnoses and measurement of the impact on public health ^[5]^. Quadriceps muscle weakness may also contribute to knee pain ^[6]^.

The quadriceps femoris muscle works as a key dynamic stabilizer of the knee, acting by virtue of its eccentric contraction to offer marked shock absorption for the knee during gait across all terrains ^[7]^. Quadriceps weakness is described as one of the earliest clinical findings, appearing prior to patient-reported symptoms ^[8-11]^. Quadriceps strength is reduced in patients in the later stages of knee osteoarthritis, indicating its role in disease evolution ^[12,13,14]^. Inadequate attenuation of forces due to quadriceps weakness and inactivity precipitates compressive forces at the knee ^[15]^. Thus, quadriceps weakness may make the knee susceptible to injury as well as exacerbate existing injury ^[3]^.

This study is a step towards analyzing the particular link that quadriceps weakness forms with symptomatic and radiographic knee osteoarthritis. Evidence that quadriceps strength progressively reduces as radiological and symptomatic grades of osteoarthritis increase may validate the theory of strengthening the quadriceps muscle to alter the disease’s course or, at the least, reduce the pain and disability associated with this severely incapacitating disease.

This study aimed to determine the isometric quadriceps strength in subjects with symptomatic osteoarthritis of the knee, to identify the association of quadriceps strength with pain and disability in subjects with knee osteoarthritis, and to compare the quadriceps strength in subjects with mild, moderate, and severe radiological knee osteoarthritis.

## METHODS

This cross-sectional study was done using the convenience sampling method over two months, involving subjects visiting the Orthopedic Outpatient Department with complaints of pain in their knee joint.

### Inclusion criteria

1. Subjects aged 40 years and above
2. Subjects complaining of pain in the knee joint, insidious in onset and having lasted longer than one month
3. Subjects having radiological osteoarthritic features as per Kellgren-Lawrence (K/L) grading with a grade more than or equal to K/L grade I

### Exclusion criteria

1. Those who are unwilling to participate in the study and are unwilling to give the required consent form
2. Subjects with a documented history of recent or old trauma to the currently painful knee, directly or indirectly causing severe bony or ligament injuries
3. Subjects with multiple arthralgias, signifying an inflammatory cause of arthritis like rheumatoid arthritis, ankylosing spondylitis
4. Subjects having undergone surgery on the currently painful knee or with a history of infection of the joint, indicating septic arthritis

Using the Western Ontario McMaster Universities Osteoarthritis Index (WOMAC), all subjects included were scored for their pain and disability ^[6]^. Subjects were also evaluated clinically for swelling and deformity. Radiographic evaluations consisted of an anteroposterior radiograph of the subject’s knee in the standing position and subsequent grading as per the Kellgren and Lawrence (K/L) system. The patients were sub-grouped into those with Mild osteoarthritis (K/L Grade I), Moderate osteoarthritis (K/L Grade II), and Severe osteoarthritis (K/L Grades III and IV) ^[6]^. Radiographs were reported by an academically based bone and joint radiologist.

A digital hand-held dynamometer manufactured by Camry in the USA was used to assess isometric quadriceps muscle strength. It was modified with straps tied onto the affected leg over the ankle to support the testing leg. The subjects were guided to sit on a fixed chair with their hips and knees flexed to 90°. They placed both hands on the distal parts of their thighs to avoid compensation. The subjects were instructed to extend the testing leg against the dynamometer’s resistance, leading to the contraction of the quadriceps muscle in an isometric manner. (**Figure 1)**

**Figure 1.**
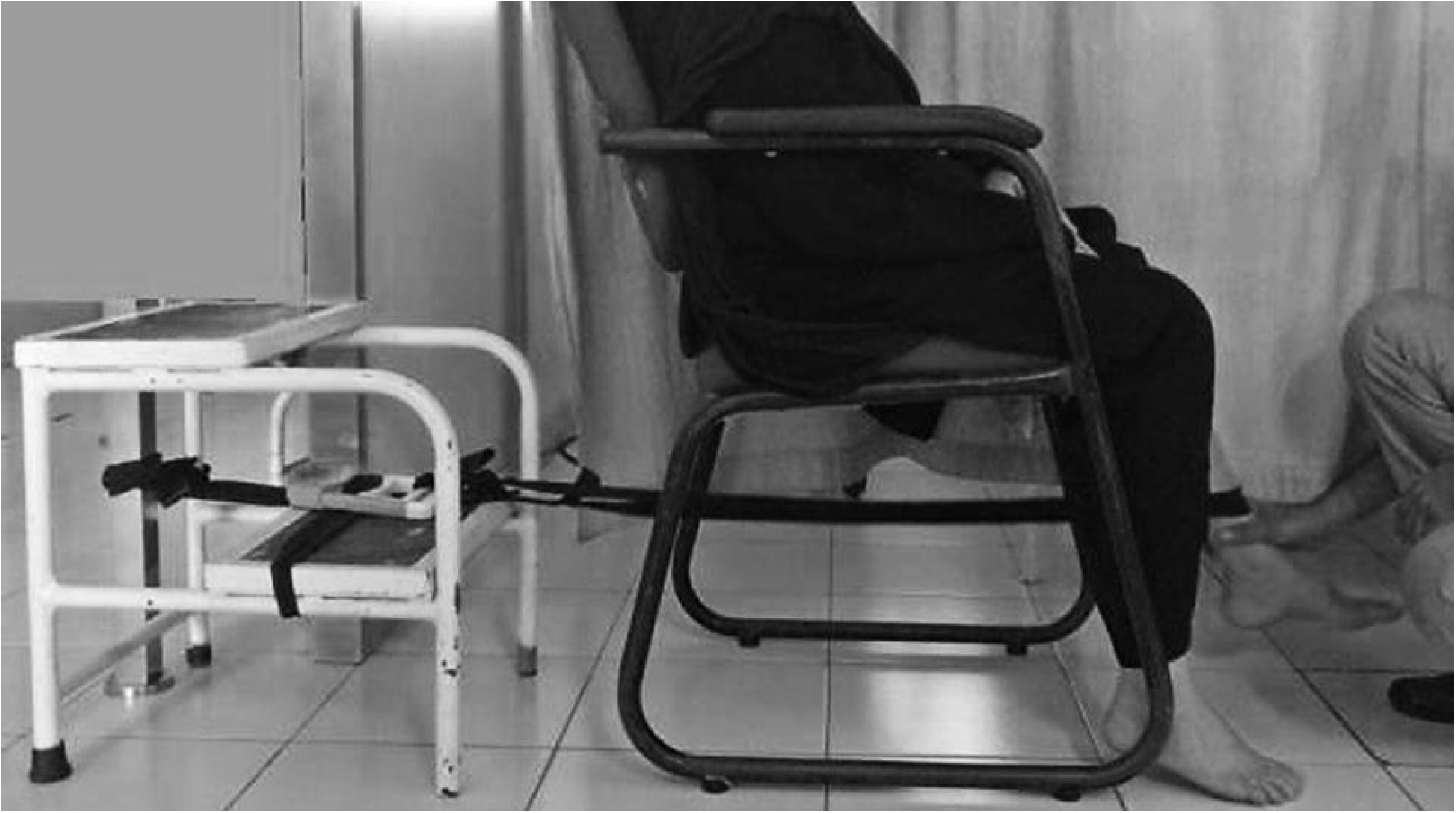
Position of the subject during the assessment of quadriceps muscle strength (with the modified hand-held dynamometer with straps)

Each patient performed three trials, with a resting time of 5 minutes between each trial, the maximum reading amongst the three trials being taken as the final reading ^[16]^. The subjects completed all assessments on the same day.

The data was entered in Microsoft Office Excel 2007, and analysis was done using IBM SPSS version-17. The comparison of the quadriceps strength in patients was made using student t-test and Pearson correlation.

## RESULTS

A total of 50 patients were included in this study, consisting of 21 males and 29 females. Females more likely had osteoarthritis **(Table 1)**. The common age group of females involved is between 55-59 years, with a mean age of 60. In males, osteoarthritis is most common between 60-65 years with a mean age of 59 years. Both the right and left knee joints are equally prone (25 subjects each) to osteoarthritic changes in both sexes. On clinical assessment, 27 subjects (54%) had no deformity, 20 subjects (40%) had mild varus deformity, 2 subjects (4%) had moderate varus deformity, and one subject (2%) had a valgus deformity. 9 subjects (18%) had no swelling, 35 subjects (70%) had mild swelling, and 6 subjects (12%) had moderate swelling in their knee. 44 subjects (88%) reported joint tenderness, and joint crepitus was eligible in 29 subjects (58%). Maximum subjects had quadriceps strength between 8-12.9 kg (34%), followed by 3-7.9kg (30%), 13-17.9kg (28%) and 18-21.9kg (8%). There is a significant difference in duration of swelling among male and female subjects **(Table 2)**.

**TABLE 1:** Stratification of study population by age and sex.

| **S.No.** | **Age (in years)** | **Males** | **%** | **Females** | **%** |
| --- | --- | --- | --- | --- | --- |
| 1. | 40-44 | 3 | 6 | 1 | 2 |
| 2. | 45-49 | 3 | 6 | 2 | 4 |
| 3. | 50-54 | 1 | 2 | 3 | 6 |
| 4. | 55-59 | 2 | 4 | 7 | 14 |
| 5. | 60-64 | 5 | 10 | 6 | 12 |
| 6. | 65-69 | 1 | 2 | 5 | 10 |
| 7. | 70-74 | 3 | 6 | 5 | 10 |
| 8. | 75-79 | 3 | 6 | 0 | 0 |
| **Total** |  | 21 | 42 | 29 | 58 |

**TABLE 2:** Difference in the duration of pain & swelling among male & female.

| **S.No.** | **Gender** | **Duration of pain (in months)** | **Duration of swelling**<br>**(in months)** |
| --- | --- | --- | --- |
| 1. | Male | 20.90±18.53 | 8.90±15.97 |
| 2. | Female | 30.24±30.34 | 2.96±1.88 |
| **t value** |  | 1.2500 | 1.9918 |
| **p-value** |  | 0.2174 | **0.0521** |

**Table 3** shows that maximum subjects with moderate osteoarthritis had quadriceps muscle strength between 13 -17.9% with a mean of 12.81±3.48 kg. Maximum subjects with severe osteoarthritis had quadriceps muscle strength between 3-7.9% with a mean of 10.96±6.29. **Table 4** indicates the quadriceps muscle strength across advancing radiological grades of knee osteoarthritis.

**TABLE 3:** Comparison of Quadriceps Strength in subjects subgrouped by K/L grades.

| **K/L Grades** | **Quadriceps Strength (in kg)** |  |  |  |  |  |  |  | **Number**<br>**of subjects** | **%** |
| --- | --- | --- | --- | --- | --- | --- | --- | --- | --- | --- |
|  | **3-7.9** | **%** | **8-12.9** | **%** | **13-**<br>**17.9** | **%** | **18-21.9** | **%** |  |  |
| Mild<br>osteoarthritis | 0 | 0 | 0 | 0 | 0 | 0 | 0 | 0 | 0 | 0 |
| Moderate<br>osteoarthritis | 2 | 4 | 6 | 12 | 9 | 18 | 1 | 2 | 18 | 36 |
| Severe<br>osteoarthritis | 13 | 26 | 11 | 22 | 5 | 10 | 3 | 6 | 32 | 64 |
| **Total** | 15 | 30 | 17 | 34 | 14 | 28 | 4 | 8 | 50 | 100 |

**TABLE 4:** Mean Quadriceps Strength in subjects subgrouped by K/L grades.

| **Number**<br>**of subjects** | **K/L grade** | **Mean Quadriceps**<br>**Strength (in kg)** | **t value** | **p value** |
| --- | --- | --- | --- | --- |
| 0 | Mild osteoarthritis | 0 | 1.1867 | 0.2412 |
| 18 | Moderate osteoarthritis | 12.81 ± 3.48 | | |
| 32 | Severe<br>osteoarthritis | 10.96 ± 6.29 | | |

A significant negative correlation exists between K/L grade and maximum quadriceps muscle strength (Q_max_), r **(50) = -0**.**28** (**p= 0**.**0489**). A highly significant negative correlation exists between the WOMAC score and strength of the quadriceps muscle (Q_max_), **r(50)= -0**.**41** (**p=0**.**0031**). **(Figure 2)**

**Figure 2.**
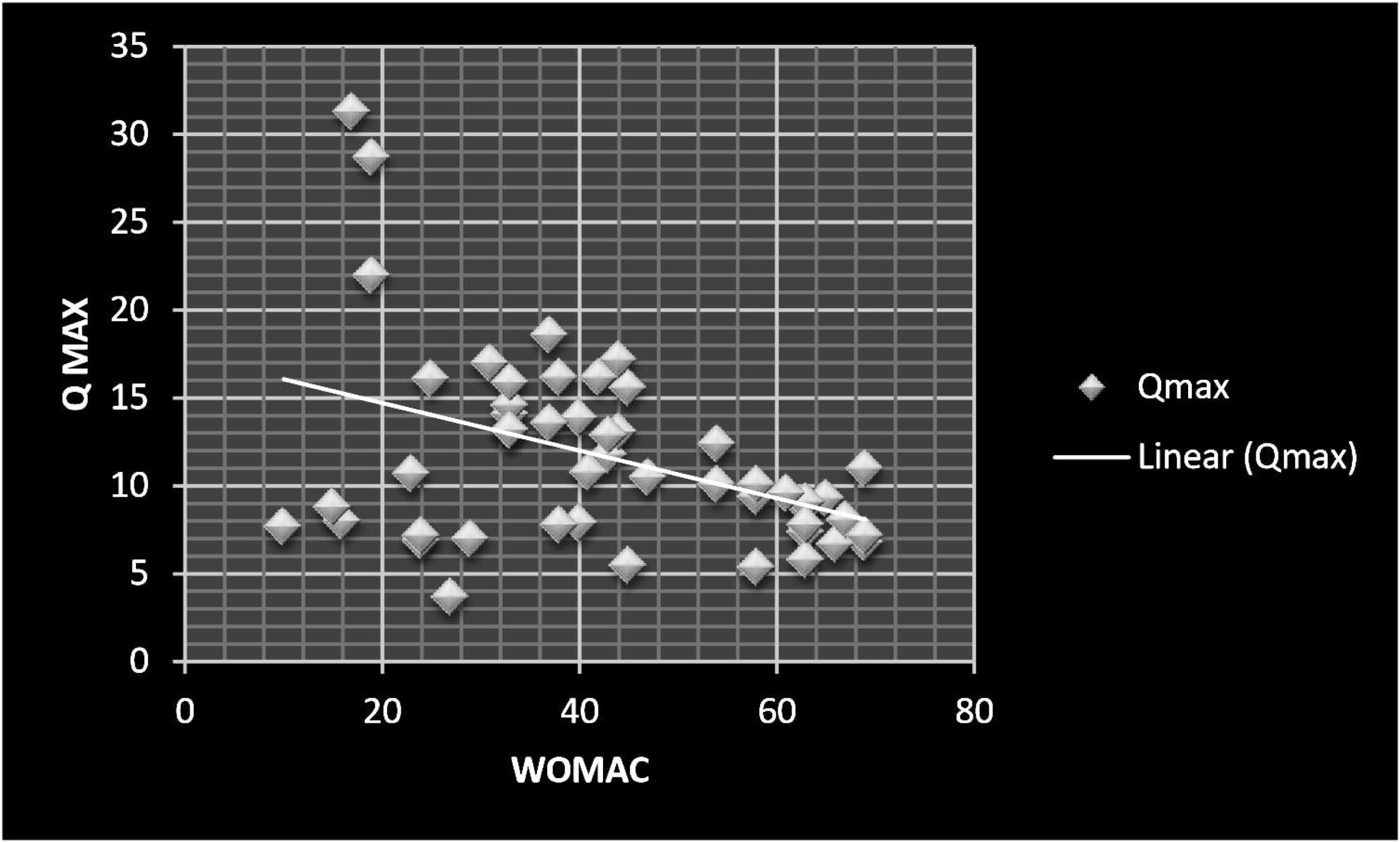
The chart shows the distribution of WOMAC scores and the negative correlation between WOMAC score and Quadriceps muscle strength (Q_max_).

## DISCUSSION

Subjects with weak quadriceps had significantly more severe symptoms and functional impairment as assessed by WOMAC scoring (p<0.05). In addition, they had significantly more structural deterioration in the knee joints as assessed by K/L grading (p<0.05).

Many cross-sectional studies support that lower quadriceps strength contributes to poor subject reported function, worse physical performance, and disease progression ^[12, 17, 18]^. Recent longitudinal data has shown that greater baseline quadriceps strength may protect against incident knee pain ^[19, 20]^. Luc-Harkey et al. found that weak quadriceps contribute to various clinical features of osteoarthritis and functional limitations in the affected knee joints ^[21]^. The declining quadriceps strength in progressive stages of knee osteoarthritis and its effect on patient-reported symptoms, both subjective and objective, and physical disability has been well established ^[6, 14, 18, 22]^.

The present study observed that the subjects with weak quadriceps had higher disability scores and marked damage on knee joint radiographs. This association questions whether the quadriceps muscle weakness precedes the onset of osteoarthritis disease and progressively results in further ongoing damage OR the joint disease itself reduces the quadriceps muscle strength. One theory states that quadriceps strength reduces in osteoarthritic knees partly due to muscle atrophy and partly due to arthrogenic muscle inhibition. These authors have described the gamma loop (γ), a spinal reflex circuit brought into play by the loss of sensory output from damaged mechanoreceptors within the osteoarthritic knee joint. This gamma loop initiates the arthrogenic muscle inhibition leading to quadriceps weakness and atrophy ^[23]^.

Palmiere-Smith et al. compared the isometric quadriceps muscle strength in women with mild, moderate, and severe radiological knee osteoarthritis. They found that women with K/L grades of 0 to 1 had stronger quadriceps than women with K/L grades of 2 to 3 and 3 to 4 ^[24]^. They concluded that quadriceps weakness is present in the later and advanced stages of osteoarthritis and present earlier in the disease process as well when the disease is classified as mild according to radiographs and MRI. Similarly, in their 30-month longitudinal study, authors Segal et al. concluded that lower quadriceps strength was associated with increased incident radiographic knee osteoarthritis in women, but not in men ^[19]^.

The present study found a significant difference in quadriceps strength between subjects with K/L grades 2 to 3 and 3 to 4. Moreover, the negative correlation between quadriceps strength and radiographic joint destruction was not as strong as the association observed between quadriceps strength and disability.

Amin et al. found no association between quadriceps strength and cartilage loss at the tibiofemoral joint. However, greater quadriceps strength was protective against cartilage loss at the lateral compartment of the patellofemoral joint ^[20]^. Other similar studies like those of Brandt et al. ^[25]^ and Thorstensson C et al. ^[26]^ found no difference in quadriceps strength between women with and without tibiofemoral osteoarthritis. This difference in the associations of quadriceps strength with incident symptomatic osteoarthritis and incident radiological osteoarthritis may be due to pain acting as a confounding variable during the assessment of the quadriceps muscle strength ^[27]^. Subjects with severe osteoarthritis are already in pain, which may aggravate during the quadriceps strength assessment. The pain elicited during the manoeuvre might reduce the maximal quadriceps contraction, and hence, the resultant muscle strength reading will be lower. This theory is substantiated by a 2×2 factorial design study by Hall M, where four subgroups were compared to normal subjects. Their study confirmed that individuals with both radiological osteoarthritis and knee pain experienced greater disability than with either radiological osteoarthritis or pain alone ^[28]^.

A modified hand-held dynamometer was used to assess the quadriceps muscle strength in subjects with symptomatic knee osteoarthritis in the present study. It is a cost-effective tool and can be easily adapted in clinical settings to provide a quick estimate of subjects’ isometric quadriceps muscle strength. Mentiplay BF et al. have substantiated the excellent reliability and validity of hand-held dynamometer in assessing lower limb muscle strength ^[29]^, which is supported by Kim SG et al., who determined the inter- and intra-rated reliability of the hand-held dynamometer ^[30]^.

In the present study, a few subjects, despite their elderly age and higher K/L grade (i.e., with more radiological destruction of knee joint), had less pain and symptoms. These subjects had better isometric quadriceps strength. A longitudinal study done to evaluate this group of subjects may demonstrate a stronger link between quadriceps strength and the development of knee osteoarthritis. As muscle strength is modifiable, appropriate measures to strengthen the quadriceps muscles may reduce the development and progression.

## LIMITATIONS

Although the present study included K/L grade 0 to 1 as mild osteoarthritis, there were no subjects identified under this group as this indicates early disease and is mostly diagnosed with the help of MRI. Furthermore, this group of subjects may not have had many symptoms to visit the hospital. As it was only a cross-sectional study, there was no follow up to assess the quadriceps strength or disease progression. Comments about causality cannot be made. Involvement of normal healthy people (control group) with matched variables of age, sex, BMI may provide more significant and valid results.

## CONCLUSION

Subjects with greater quadriceps muscle strength had lower disability scores and lesser structural damage. With the progress of the osteoarthritic changes, there is a gradual reduction in quadriceps muscle strength. Our study established a stronger association between quadriceps strength and functional disability, indicating that even though subjects had higher or greater joint destruction on knee radiographs, increasing the strength of quadriceps might reduce their pain and functional disability. Any physical interventions to strengthen the quadriceps at an appropriate stage will reduce the disease’s progression and prevent a worse functional outcome.

## Data Availability

The data obtained and analyzed during the current study are available from the corresponding author on reasonable request.

## Acknowledgements

The authors thank Indian Council of Medical Research (ICMR) for involving the corresponding author in its Short Term Studentship (STS) Programme.

## ABBREVIATIONS

BMI: Body Mass Index
K/L: Kellgren and Lawrence
MRI: Magnetic Resonance Imaging
USA: United States of America
WOMAC: Western Ontario McMaster Universities Osteoarthritis Index

